# Profiling Neuronal Methylome and Hydroxymethylome of Opioid Use Disorder in the Human Orbitofrontal Cortex

**DOI:** 10.1101/2022.09.09.22279769

**Authors:** Gregory Rompala, Sheila T. Nagamatsu, José Jaime Martínez-Magaña, Jiawei Wang, Matthew J. Girgenti, John H. Krystal, Joel Gelernter, Traumatic Stress Brain Research Group, Yasmin L. Hurd, Janitza L. Montalvo-Ortiz

## Abstract

Opioid use disorder (OUD) is influenced by genetic and environmental factors. While recent research suggests that there are epigenetic disturbances in OUD, these studies were limited to DNA methylation (5mC). DNA hydroxymethylation (5hmC) has been understudied in psychiatric disorders and addiction, despite being highly enriched in the brain where it regulates critical functions, including neural plasticity. Here, we conducted a multi-omic profiling of the orbitofrontal cortex (OFC) of OUD, integrating neuronal-specific 5mC and 5hmC as well as within-subject correlations with gene expression profiles from human postmortem samples (OUD=12; controls=26). Further, co-methylation modules and GWAS enrichment were analyzed for 5mC and 5hmC. Evaluation of single locus methylomic alterations identified 397 and 1740 differentially 5mC and 5hmC CpGs, respectively. Enrichment for neuronal function was observed for 5hmC, while no significant pathways were observed for 5mC. 5mC and 5hmC co-methylation analysis identified modules associated with OUD enriched for Pre-NOTCH Transcription and Translation, and WNT signaling. Transcriptomic analysis identified *HBB* as significantly associated with OUD. Finally, drug interaction analysis showed seven differential 5hmC genes and one differential 5mC gene interacting with opioid use. Our multi-omic findings suggest an important role of 5hmC and reveal novel loci epigenetically dysregulated in OFC neurons of individuals with OUD.

## Background

Opioid use disorder (OUD) is a serious public health problem because of its high disease burden, as measured by hospitalization and drug overdose death rates^1^, and involvement in the criminal justice system based on the high incarceration rates^2^. In the U.S., opioid overdose deaths reach 17.8 per 100,000 individuals. This opioid epidemic worsened during the COVID-19 pandemic, as shown by a significant steep rise of 29.4% in opioid overdose deaths in 2020^3^.

OUD is associated with a wide range of acute effects and longer-term brain neuroadaptations related to intoxication, tolerance, and dependence and which can contribute to compulsive opioid use^4, 5^. Activation of µ, ∂, and k opioid receptors alter the activity of stress and reward circuitry. In animals and humans, the orbitofrontal cortex (OFC) has been implicated in the development and maintenance of drug addiction, deficits in inhibition of impulsive behavior, and distortions in reward-related decision-making processes^6, 7, 8, 9^.

Multiple kinds of epigenetic modifications regulate gene expression that impacts behaviors relevant to opioid addiction^10^. Changes in DNA methylation at CpG dinucleotides (5mC) are associated with OUD^11^. In postmortem human OFC, heroin use was associated with differential methylation of several gene classes, including those implicated in glutamate neurotransmission, axonogenesis, synaptic processes, and the regulation of gene expression^11^. 5mC is actively oxidized to hydroxymethyl-cytosine (5hmC) by translocation (TET) enzymes, and studies have suggested that 5hmC contributes to a proper brain development^12^. 5hmC is a relatively stable epigenetic mark, highly enriched in neurons and associated with promoting transcription^13^. Genome-wide differential 5hmC studies in human postmortem brain have suggested an association with antemortem psychiatric traits, including depression^14^ and alcohol use disorders^15^. Although there are no prior studies of 5hmC in human OUD, a study in rodents chronically exposed to morphine reported changes in both 5mC and 5hmC levels (global and promoter-specific) across multiple brain regions^16^.

To date, studies of epigenetic changes in postmortem human brain tissue have been limited to analyses of bulk tissue samples featuring diverse neuronal and non-neuronal cell-types. As epigenetic marks regulate gene expression in a cell-type-specific manner, the mixing of cell types in bulk tissue analyses may obscure cell-type-specific findings.

Here, we conducted the first parallel 5mC and 5hmC profiling of OUD in neuronal nuclei from human postmortem OFC. Further, we integrated gene expression data to elucidate the effect of differential 5mC and 5hmC on gene transcription. Lastly, we performed a GWAS enrichment analysis to examine whether differential 5mC and 5hmC marks are related to genetic variation associated with OUD. Our integrative multi-omic study uncovered novel neuronal 5mC and 5hmC functional marks and co-methylation networks associated with OUD in the human OFC.

## Results

### Demographics and clinical characteristics

Brain samples from the OFC included 12 OUD cases and 26 non-OUD controls (**Table 1**). All brain samples were from males of European and African ancestry. There was no significant difference in the age of death between OUD (mean=43.1; SD=11.55) and non-OUD groups (mean =37.6; SD=8.9), as well as post-traumatic stress disorder (PTSD) (n=12, p-value=0.84), major depressive disorder (MDD) (n=8, p-value=0.40), and cigarette smoking (n=10, p-value=0.53).

**Table 1:** Demographics Summary.

|  | OUD- (N=26) | OUD+ (N=12) |
| --- | --- | --- |
| Age ($\mu$ +/- SD) | 43.1 +/- 11.55 | 37.6 +/- 8.9 |
| PMI ($\mu$ +/- SD) | 30.65 +/- 8.15 | 29.6 +/- 7.5 |
| Ancestry |  |  |
| AA | 5 | 4 |
| EA | 19 | 8 |
| Cigarette Smoking | 13 | 10 |
| Alcohol Dependence | 5 | 3 |
| PTSD | 13 | 12 |
| Major Depressive Disorder | 5 | 8 |
| Polysubstance Abuse | 4 | 7 |
| Removed Outliers | 2 | 0 |

### 5mC and 5hmC assessment in OFC Neurons

We used fluorescence-activated nuclei sorting (FANS) with the postmitotic neuronal marker NeuN and reduced representation oxidative bisulfite sequencing (RRoxBS) to profile 5mC and 5hmC in OFC neuronal nuclei isolated from postmortem brain samples (Figure 1A, **Supplementary Figure 1**). All CpG sites (CpGs) with ≥10 base pair coverage were analyzed, with most CpGs at 40-60X coverage (**Figure 1B**). An average of 10X coverage was obtained for ~3.5 million CpGs. Overall, we analyzed 1,842,090 CpGs for 5mC and 1,653,870 for 5hmC. Principal component analysis of all 5mC and 5hmC samples identified two outlying subjects removed from the final analysis (**Supplementary Figure 2**). The greatest percentage of CpGs (44%) were in gene promoter regions (+/-1 kb from the transcription start sites - TSS), followed by intergenic (30%), intronic (22%), and exonic regions (4%) (**Figure 1C**). 5mC and 5hmC were significantly reduced in promoter and intron regions of neuronal marker genes compared to oligodendrocyte markers, although 5hmC was increased in exons for neuronal genes (**Figures 1D, 1E**). In addition, we analyzed previously established differentially methylated regions in cortical neuronal nuclei^17^, and show a strong relationship between total methylation (5mC+5hmC) at CpGs in neurons of that dataset and neuronal OFC total methylation of the same CpGs in this study (R^2^=0.66, p<0.0001; Figure 1F). Similarly, CpG methylation in OFC NeuN+ neurons was inversely correlated with CpG methylation previously observed in cortical non-neuronal nuclei (R^2^ =-0.44, p<0.0001; **Figure 1F**).

**Figure 1.**
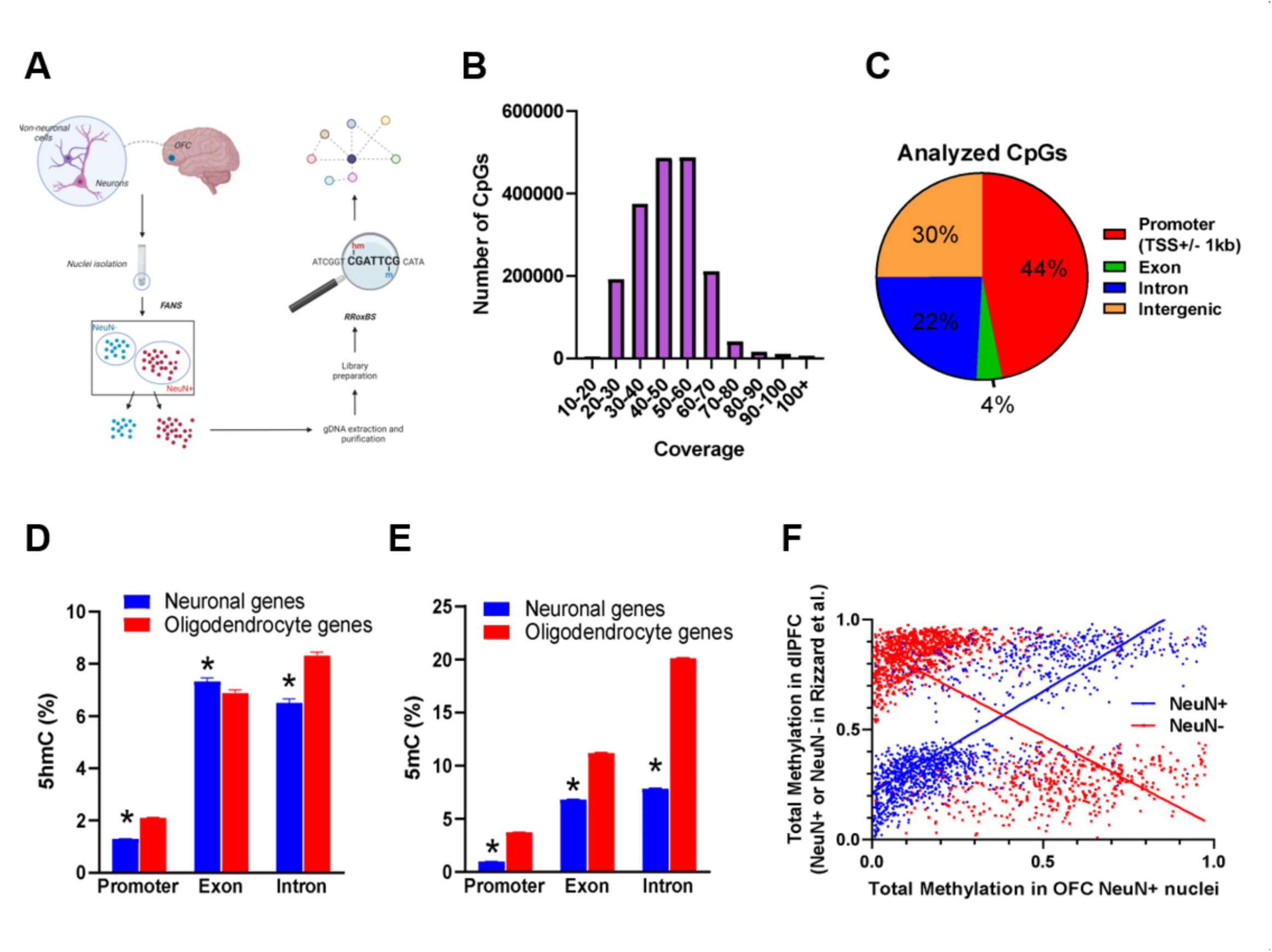
Analyzing the Neuronal Methylome and Hydroxymethylome in OUD. A) Experimental workflow. Fluorescence-Activated Nuclei Sorting was used to isolate neuronal nuclei from postmortem OFC. Nuclei were processed for genomic DNA, undergoing reduced representation oxidative bisulfite sequencing to examine 5mC and 5hmC at CpG-dense loci. (B) An average of 10x coverage was obtained for ~3.5 million CpG sites. C) 44% of CpG sites were located in promoter regions and 30% in intergenic regions. D-E) Neuronal 5mC occurs mainly in intergenic regions, while 5hmC occurs in introns, exons, and intergenic regions. F) Contrasting mean 5mC levels at neuronal (NeuN+) vs. non-neuronal (NeuN-) marker genes.

### Differential 5mC and 5hmC associated with OUD

Individual CpGs were evaluated for differential 5mC and 5hmC levels in OUD vs. non-OUD subjects. For 5mC, we found 397 differential CpGs (357 genes) (**Supplementary Table 1**); for 5hmC, we identified 1,740 differential CpGs (1,453 genes) (**Supplementary Table 2**). There was no overlap between differentially 5mC and 5hmC marks. Considering the gene nearest to each differential 5mC or 5hmC in OUD, 56 genes had at least one differential 5mC and 5hmC mark (15.4% of 5mC-linked genes and 3.5% of 5hmC-linked genes) (**Figures 2A and 2B**). Evaluating differential CpGs across genomic loci indicated that most 5mC and 5hmC CpGs were located in promoter regions within 1 kb of a TSS. (**Figure 2C**). Indeed, examining genomic regions with known associations with the histone modification H3K27ac indicates these are likely active promoter and enhancer regions^18^. Moreover, the enrichment was more significant in neuron-specific vs. non-neuronal-specific H3K27ac regions (**Figure 2D, Supplementary Table 3**). Differential 5mC marks were most significantly enriched in GABAergic neuronal marker genes, and differentially 5hmC CpGs were significantly enriched for neuronal marker genes (i.e., genes increased in both GABAergic and glutamatergic neurons vs non-neuronal cell types) (**Figure 2E, Supplementary Table 3**). Gene ontology (GO) enrichment analysis revealed no significant pathways among OUD-linked 5mC genes, but trending (FDR-adjusted p-value<0.1) top ontologies included nicotine- and opioid-signaling pathways (**Figure 2F, Supplementary Table 4**). Comparatively, OUD-linked 5hmC genes were significantly enriched for several terms (FDR-adjusted p-value<0.05), many related to neuronal function (e.g., G protein signaling, postsynaptic differentiation, and GABAB receptor signaling). Opioid-signaling pathways were similarly trending toward significance (FDR-adjusted p-value=0.05, **Figure 2G, Supplementary Table 5**). We compared our findings with Kozlenkov et al.^11^, who examined methylation in the postmortem OFC of heroin users (N=87) using the Illumina 450K Infinium microarray. Very few differentially methylated CpGs identified in that study had sufficient read coverage to be examined in the present study (175 of 1391). None of these sites were differential 5mC or 5hmC CpGs in OUD cases, which might be attributable to technical differences, low coverage in these CpG sites, or demographic divergences between the studies. But when comparing genes with differential CpGs and using the same significance threshold as in Kozlenkov et al. (p<0.001)^11^, we found that 256 CpGs are commonly differentially methylated between opioid overdose^7^ and OUD. The overlap increases to 327 CpGs when considering both 5mC and 5hmC (**Figure 2H, Supplementary Table 6**). GO analysis revealed that the shared genes were associated with axon guidance (5mC+5hMC; odds ratio=3.08, FDR-adjusted p-value<0.001, **Supplementary Table 7**).

**Figure 2.**
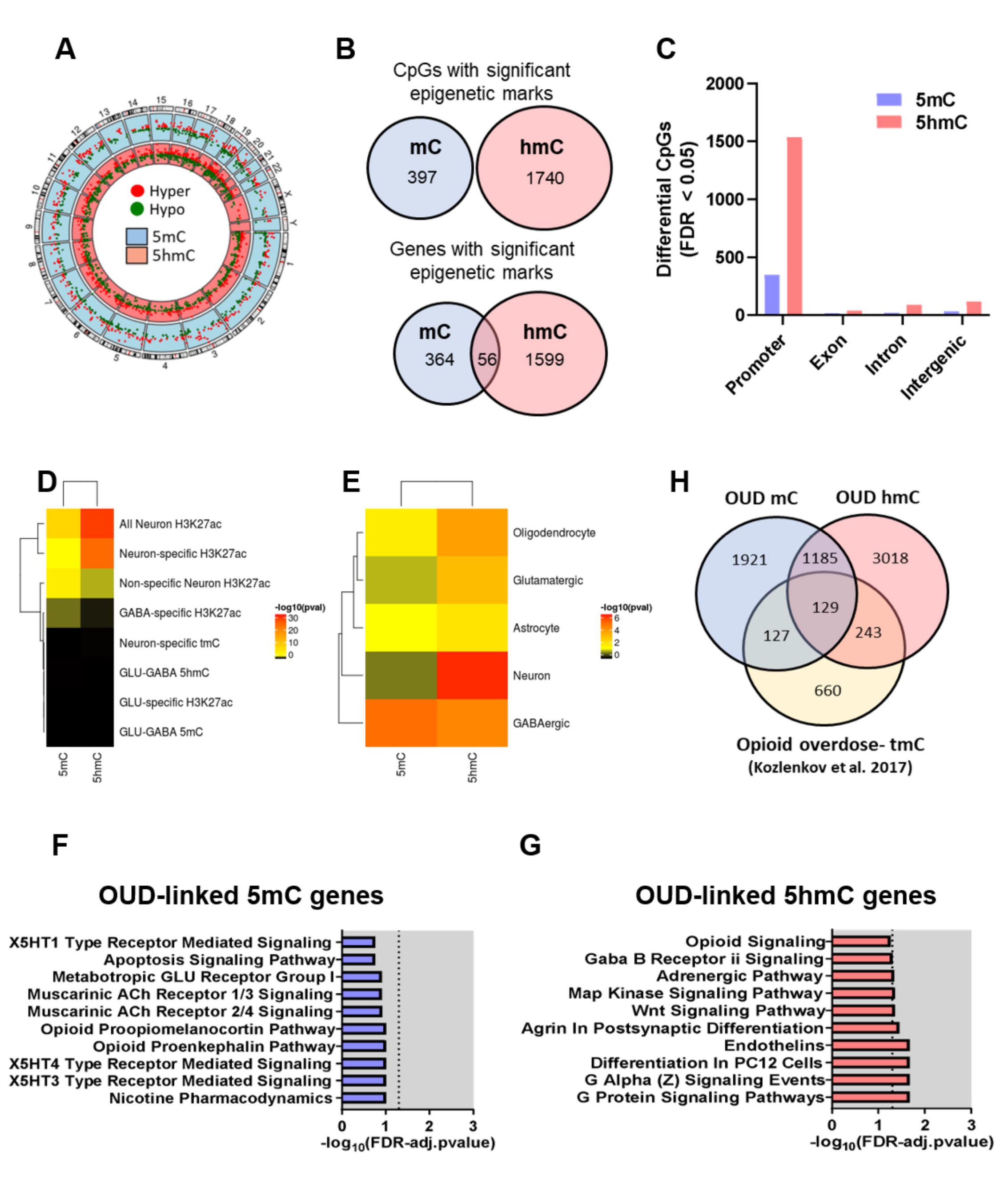
Differential 5mC and 5hmC marks of OUD. A) Distribution of differential sites into hyper- and hypomethylation for 5mC and 5hmC. B) Comparison between differential 5mC and 5hmC sites for CpG and gene. C) Distribution of differential CpGs across genomic loci. D-E) Enrichment analysis for neuronal regions. F) Comparison of differential CpGs with methylation markers detected in the prefrontal cortex of opioid overdose cases (Kozlenkov). G) Gene ontology analysis for OUD-linked 5mC genes. H) Gene ontology analysis for OUD-linked 5hmC genes.

### Gene drug interaction analyses

We evaluated predicted drug interactions for the annotated genes with differential 5mC and 5hmC CpGs and observed 616 interactions for 5mC and 2,562 for 5hmC (**Supplementary Tables 16 and 17**). We also observed interactions with 15 opiates (**Figure 6**). For the differential 5mC CpGs, we observed only one interaction for apomorphine in the gene *CALY*. For the 5hmC, we observed a predicted interaction of opioids with 7 genes including *CBFB, GRIN1, HCN1, HMOX2, MPO, RUNX1*, and *SOD2*. From all 5hmC genes, only *HCN1* showed interaction only with opioids (tramadol). *MPO* also interacts with additional drugs, including anti-inflammatory drugs (i.e., nimesulide, tolmetin, and diclofenac). *GRIN1* interacts also with glutamate receptor (i.e., dizocilpine, (d)-serine), and pain management (ketamine, ralfinamide, orphenadrine, orphenadrine citrate) drugs. *CBFB* interacts with a dopamine receptor agonist drug, ergocornine.

### Co-methylation analysis of OUD

In addition to EWAS, we also conducted a co-methylation analysis using WGCNA, a weighted gene co-expression networking analysis that allows us to identify clusters of CpG sites (modules) with similar methylation levels. By applying this method, we aimed to assess the intercorrelation among CpG sites and identify epigenetically-defined gene networks associated with OUD. This analytical approach to assess co-methylation has been previously applied in the context of alcohol use disorder in the human postmortem brain^19^. Here, co-methylation analysis identified 626 modules for 5mC and 572 for 5hmC. We analyzed module eigengene associated with OUD (cor>|±0.4| and p-value<0.05), including ten modules for 5mC (**Figure 3A**) and four modules for 5hmC (**Figure 3B**). Module membership vs. gene significance is shown in **Supplementary Figure 3**. For 5mC, six modules showed enrichment for GO terms (**Supplementary Table 8**), of which the top 10 are shown in **Figure 3C**. The 5mC module with the strongest correlation with OUD (cor=0.48) was the Steelblue1 enriched for the Pre-NOTCH Transcription and Translation (p-value=5.71E-03; **Supplementary Table 10**. Most of the OUD-associated 5mC modules were enriched for transcription regulation, cell differentiation, nervous system development, morphogenesis, and generation of neurons. Further, two of those modules, turquoise3 and lightpink2, we observed enrichment for the following Reactome pathways: Inhibition of Voltage-Gated Ca^2+^ Channels via Gbeta/gamma Subunits, Activation of GABAB Receptors (p-value range: 1.44E-02 to 6.55E-03, turquoise3), angiotensin II-stimulated signaling through G proteins and beta-arrestin and Signaling by WNT (p-value range: 1.38E-02 to 4.44E-02, lightpink2). OUD-associated 5hmC modules were enriched for organ development, nervous system development, neurogenesis, transcription regulation, and morphogenesis (**Supplementary Table 9, Figure 3D**). Enriched Reactome pathways (**Supplementary Table 10**) included GPCR Ligand Binding (p-value=1.77E-03; thistle1), Class B/2 (Secretin Family Receptors) (p-value=7.60E-03, rosybrown1), PI3K Cascade, and Insulin Receptor Signaling Cascade (p-value range: 1.18E-02 to 4.95E-02, orange4). Protein-protein interaction (PPI) analyses were conducted for the OUD-associated 5mC and 5hmC modules (**Figure 4**) using co-expression evidence. PPI networks were enriched for several biological pathways, including regulation of biological process (GO:0050789), nitrogen compound metabolic process (GO:0006807), neurogenesis (GO:0022008), cell differentiation (GO:0030154), gene expression (GO:0010467), neuron development (GO:0048666), regulation of neurogenesis (GO:0050767), regulation of cell communication (GO:0010646), and response to stimulus (GO:0050896).

**Figure 3.**
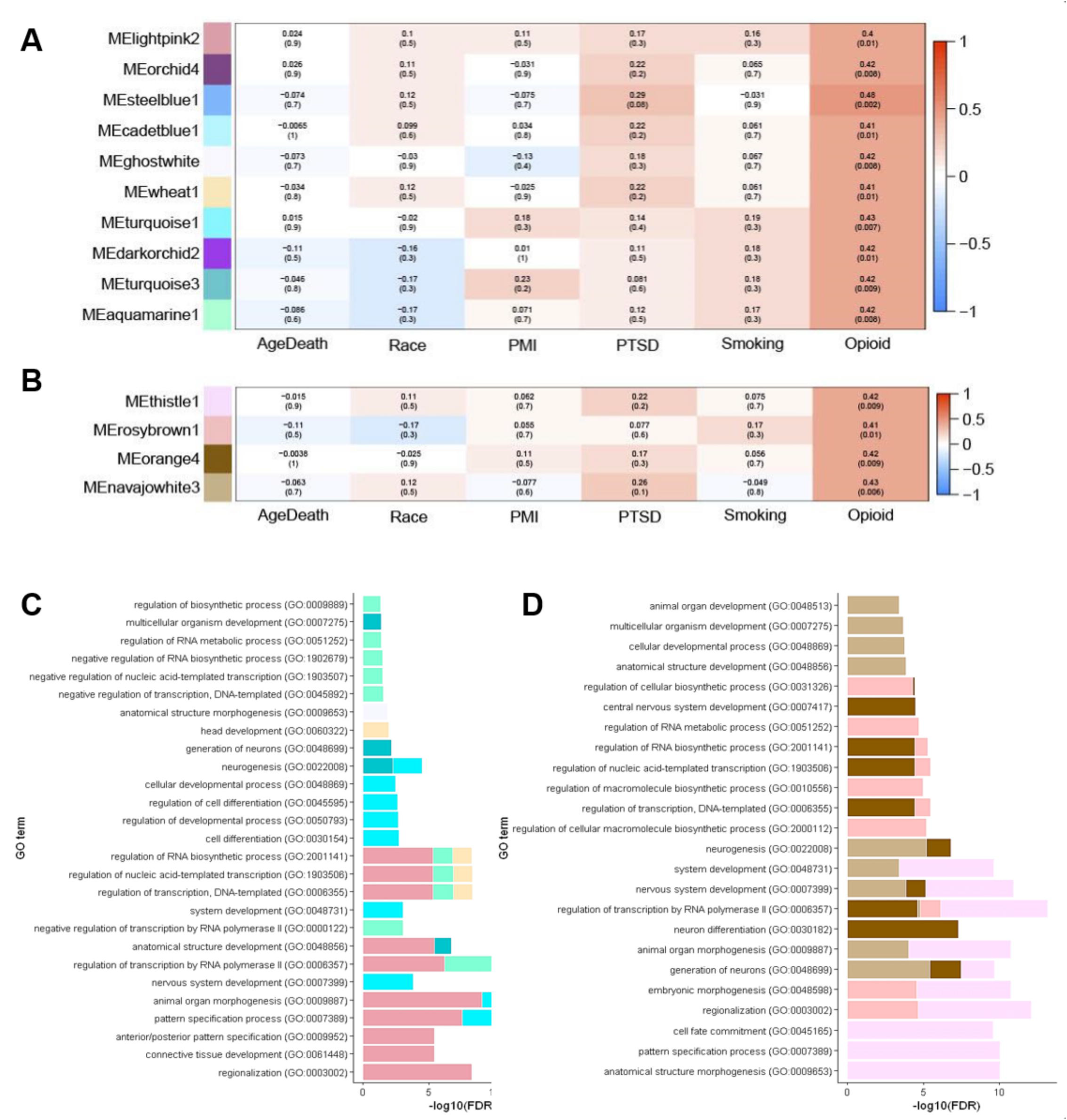
Co-methylation of 5mC and 5hmC sites. A) 5mC-significant modules associated with OUD (correlation>=|0.4|, p-value<0.05). B) 5hmC-significant modules associated with OUD (correlation>=|0.4|, p-value<0.05). C) GO enrichment analysis for 5mC-significant modules (top 10 terms). D) GO enrichment for 5hmC-significant modules (top 10 terms).

**Figure 4.**
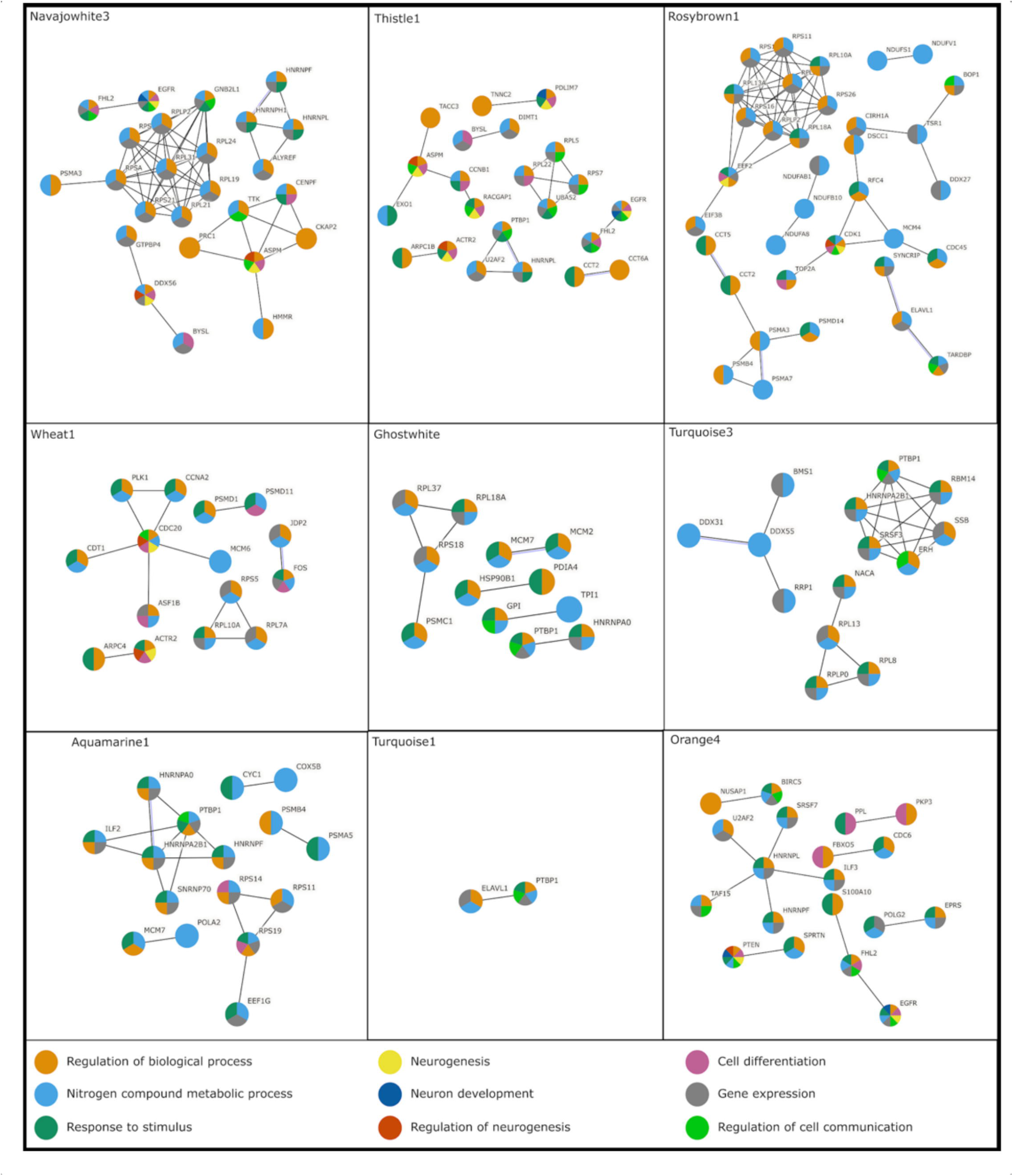
Network of 5mC- and 5hmC-significant modules associated with OUD. The figure shows the PPI analyses for 9 modules detected with a significant association with OUD in 5mC (wheat1, ghostwhite, turquoise3, aquamarine1, turquoise1) and 5hmC (navajowhite3, orange4, rosybrown1, thistle1). The colors represent a different biological process in which the modules were enriched.

**Figure 5.**
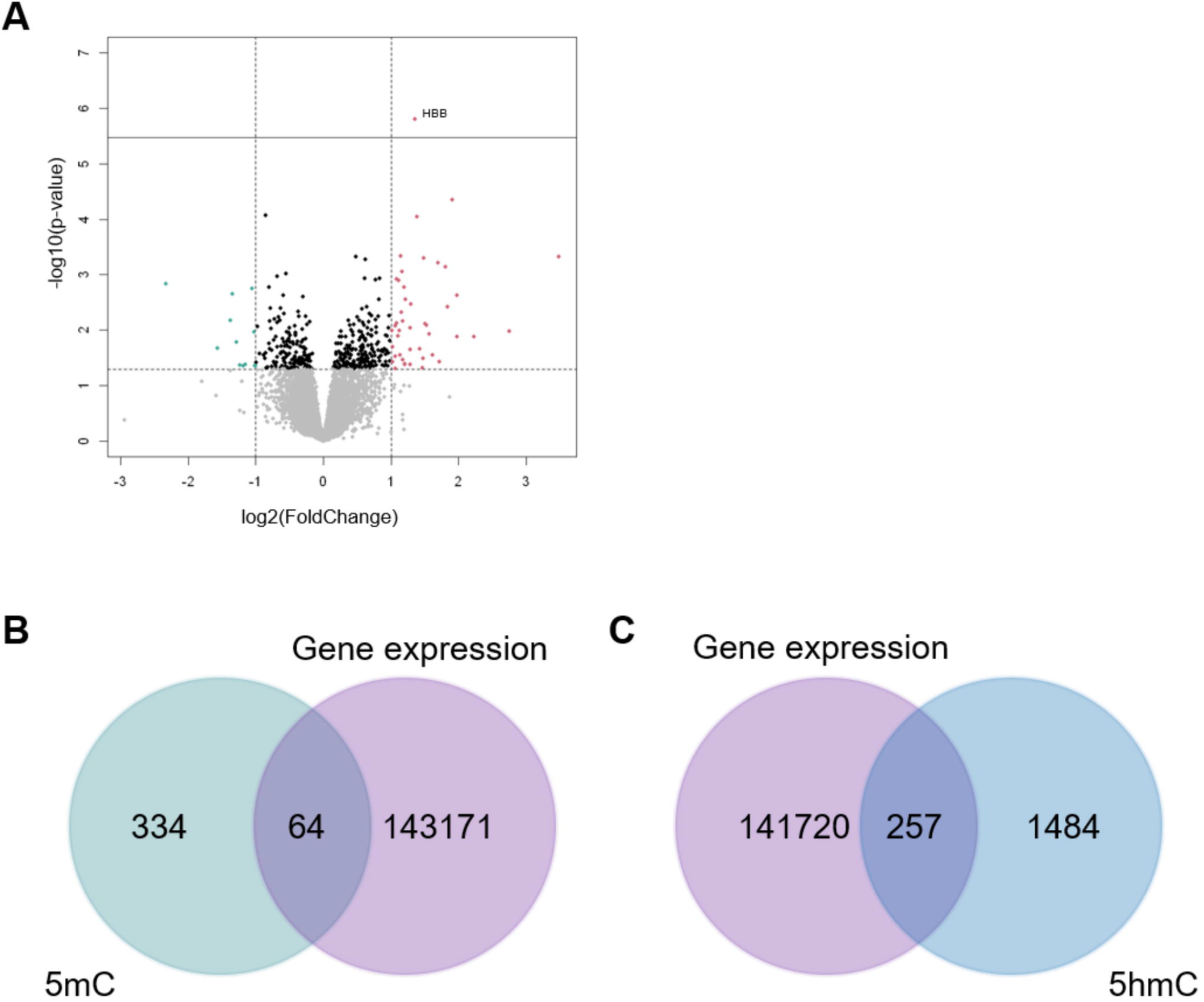
Differential expression analysis of OUD. A) Volcano plot (threshold: most significant, 10^−5^). B) Venn diagram for the methylated sites correlated with gene expression against differential 5mC CpG sites. C) Venn diagram for the hydroxymethylated sites correlated with gene expression against the differential 5hmC CpG sites.

**Figure 6.**
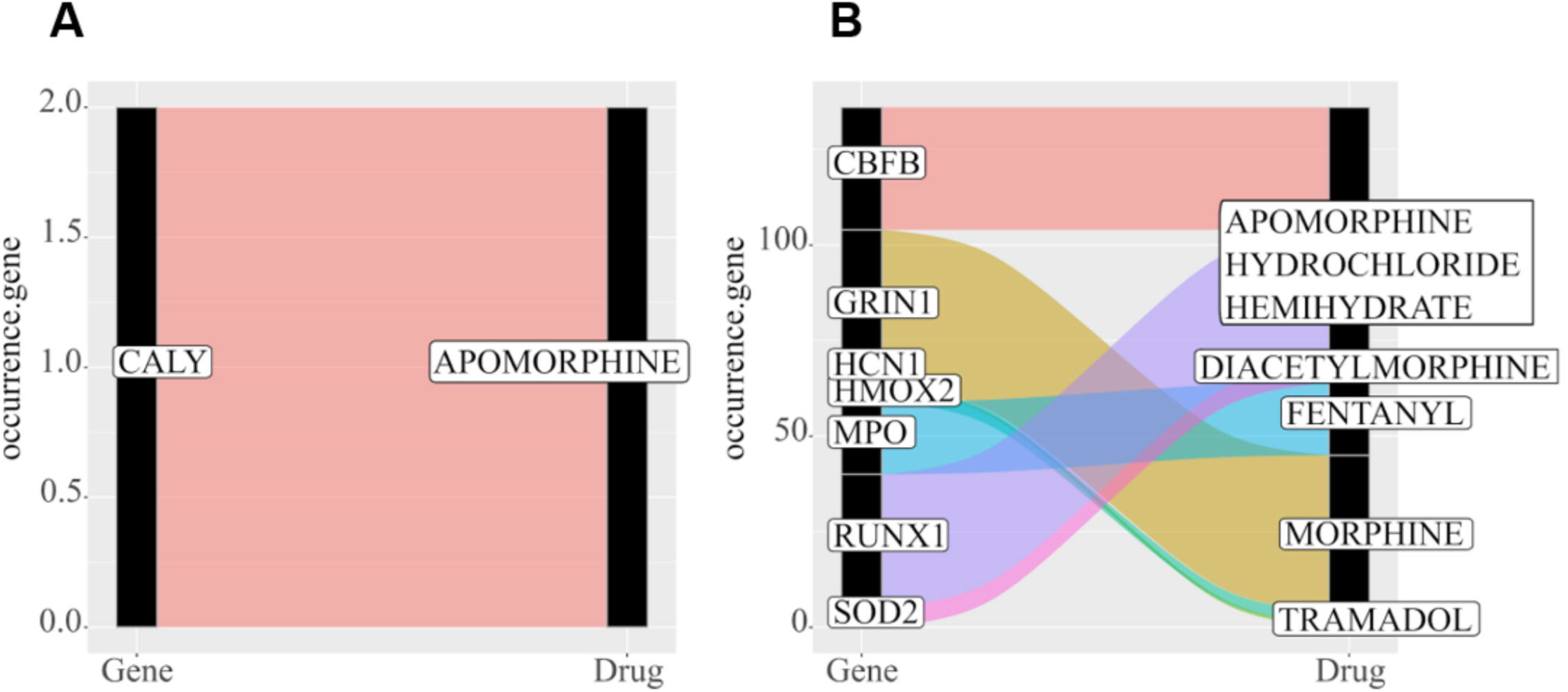
Drug interaction analysis of 5mC and 5hmC differential CpG sites with opioids. A) 5mC OUD-associated differential CpG sites. B) 5hmC OUD-associated differential CpG sites.

We also evaluated the enrichment of differential 5mC and 5hmC marks in the identified OUD-associated module networks. For the 5mC OUD-associated modules, we identified *DDX31* in the turquoise3 module. For module wheat1, we found one differential 5mC annotated gene antisense to the *JDP2* gene. No differential CpGs were enriched in the aquamarine1, ghostwhite, lightpink2, or turquoise1 modules. For the 5hmC OUD-associated modules, we identified three genes with differential 5hmC CpGs (*RPL21, RPS21*, and *ALYREF*) enriched in the navajowhite3 module. In the thistle1 module, we detected differential 5hmC CpGs in *ARPC1B, DIMT1, PTBP1*, and *CCT2*. For the rosybrown1 module, we found enrichment of differential CpGs in *RPLP2, EIF3B, CCT2, NDUFB10*, and *ELAVL1*. Lastly, we identified two genes with differential CpGs, *SRSF7*, and *POLG2*, as enriched in the orange4 module.

### GWAS enrichment analysis

We conducted a GWAS enrichment analysis to further assess the functionality of the identified differential 5mC and 5hmC genes and gene clusters. No GWAS enrichment was found for differential 5mC genes. However, differential 5hmC genes showed enrichment for skeletal domain, mainly height and structure related to movement (adj. p-value range: 4.25E-12 to 1.86E-09; **Supplementary Table 14, Supplementary Figure 5A**). We also detected GWAS enrichment in the psychiatric domain, including hyperkinetic disorders, pervasive developmental disorders, bipolar affective disorder, sleep functions, temperament and personality functions, and schizophrenia at subdomain level (adj. P-value range: 5.03e-7 to 0.044).

GWAS enrichment analysis was also performed for all 14 identified OUD-associated co-methylation modules (**Supplementary Table 15, Supplementary Figure 5B**). We detected a similar enrichment pattern to the 5hmC differential CpG sites, with higher enrichment of sites associated with the skeletal domain (adj. p-value range: 2.96E-30 to 0.018). In addition, for the 5hmC OUD-associated modules navajowhite3, orange4, thistle1, and rosybrown1, and the 5mC OUD-associated module lightpink2, we detected enrichment of genes related to the cognitive function domain (adj. p-value range: 0.00090 to 0.046). For the 5mC and 5hmC OUD-associated modules, we found enrichment in the psychiatric domain, including the following subchapter levels: sleep functions, depressive episode, schizophrenia, bipolar affective disorder, temperament and personality functions, recurrent depressive disorder, failure of genital response, hyperkinetic disorders, mental and behavioral disorders due to use of tobacco, and mental and behavioral disorders due to use of alcohol (adj. p-value range: 2.83E-19 to 0.047).

### Bulk OFC gene expression analyses

A differential gene expression (DEG) analysis was performed on 38 bulk tissue samples (OUD=12, non-OUD=26), using the same samples as in the 5mC/5hmC analyses. After Bonferroni correction, one gene, *HBB*, was differentially expressed in association with OUD (p-value=1.58E-06, **Supplementary Table 11, Figure 4A**). We also conducted a correlation analysis between bulk gene expression and neuronal-specific 5mC/5hmC levels to explore the effect of these epigenetic marks on gene regulation. We observed 64 and 257 differential 5mC (**Supplementary Table 12**) and 5hmC (**Supplementary Table 13**) CpGs, respectively, correlated with gene expression level. The two genes with the highest correlation between differential 5mC/5hmC-gene expression included the long non-coding RNA *LINC01002* and *TMIGD3*.

## Discussion

This is the first comprehensive parallel investigation of neuronal 5mC and 5hmC in the context of OUD in the human brain. Our main findings suggest an important regulatory role of 5hmC in OUD, as shown by the higher number of differential CpGs and OUD-correlated modules identified, as well as a higher concordance with gene expression patterns. Dysregulated 5mC and 5hmC CpG sites and modules were enriched for neuronal function and development and nominally associated with opioid-related signaling. Further, OUD-linked 5hmC marks were also enriched for the G protein signaling pathway and Wnt signaling as well as psychiatric domains. Drug interaction analysis indicated one gene interacting with OUD in the 5mC differential CpGs and seven in 5hmC differential CpGs including *RXN1, GRIN1*, and *CBFB*. We identified *HBB* as a differential expressed gene in OUD and observed correlations between 5mC/5hmC and gene expression patterns, suggesting a role of these epigenetic marks on gene regulation.

Most CpG sites covered by RRoxBS were located in promoter (44%) or intergenic regions (30%). Cell type enrichment analysis showed significant differences between 5mC and 5hmC CpGs, and between neuronal and non-neuronal cells. We observed reduced 5mC at neuronal-marker genes compared to oligodendrocytes, consistent with our sample being neuronal-specific. We expect a permissive 5mC signature promoting active gene expression^13^ and a more repressive 5mC signature at non-neuronal genes. Comparatively, as 5hmC in the gene-body (mainly exons) is associated with active gene expression, here we show that 5hmC levels are higher in neuronal-specific (active) genes compared to oligodendrocytes (inactive) genes. The differences observed between 5mC and 5hmC CpG sites in terms of cell-type enrichment and CpG location suggest distinct gene regulatory mechanisms among these two epigenetic modifications.

Genome-wide differential CpG analysis identified 397 CpGs for 5mC and 1,740 CpGs for 5hmC associated with OUD. These OUD-linked 5mC and 5hmC marks were primarily located in promoter regions and enriched for the H3K27ac histone modification (a marker of active enhancer), suggesting a direct role in gene regulation. Furthermore, the nearest genes annotated to both 5mC and 5hmC marks were enriched for GABAergic neuron marker genes (i.e., differentially expressed versus glutamatergic neurons. Functional analysis showed nominal enrichment in both 5mC and 5hmC for opioid-related pathways: 5mC CpGs were enriched in the opioid proenkephalin pathway and opioid proopiomelanocortin pathway and 5hmC CpGs in opioid signaling. 5hmC showed a higher number of enriched pathways, some of which have been previously implicated with OUD, including Wnt signaling^20^ and G-protein signaling. Morphine and other opioids are known to activate μ-opioid G protein coupled receptors to elicit tolerance and dependence^21^. GWAS enrichment analysis of 5hmC differential genes identified psychiatric domains, including schizophrenia and bipolar affective disorders. When evaluating the concordance of our findings with a previous study assessing 5mC in the postmortem OFC of individuals with heroin abuse who died from overdose^11^, we found some overlap when considering 5mC, 5hmC, or both. Of the 327 overlapping CpG sites, 127 were from 5mC, showing replication of our OFC OUD results.

We then evaluated gene-drug interactions for the annotated genes with differential 5mC and 5hmC and examined those with interactions with opioids. We observed that annotated genes with differential 5hmC marks showed a higher number of opioid interactions than 5mC, suggesting a higher functional role of 5hmC in OUD. *CALY* was annotated in the 5mC differential genes, and it has been previously associated with smoking initiation in a Chinese and American population^22^. In the 5hmC differential genes, we identified *GRIN1*, known to regulate μ-Opioid activity^23^; *SOD2*, previously described as associated with the heroin dependence risk in an Iranian population (n=1241)^24^, and *HCN1*, shown to be required for the activation of μ opioid receptor^25^. We also identified opioid interactions with *RUNX1*, known to bind in the promoter region of *ADRA1A* gene. Hypermethylation in the *ADRA1A* gene was previously associated with OUD in a candidate gene study in Han Chinese population ^26^. *HMOX2, MPO* and *CBFB*, also identified, have not been previously described in OUD. Lofexidine and tizanidine (drugs previously used for treatment of OUD symptoms) were observed in 5hmC interacting with *ADRA2A*, a gene involved in the release of neurotransmitters. Our results show that epigenetic marks, mostly 5hmC, functionally interact with opioids and pinpoint promising targets for OUD treatment.

In addition to differential analysis, we also conducted a co-methylation analysis to identify OUD-associated epigenetically-regulated gene networks. In the co-methylation analysis, we observed enrichment of the Reactome pathway Pre-NOTCH Transcription and Translation. NOTCH signaling has been involved in the adaptation to chronic morphine exposure and proposed as a potential target for pain management^27^. Besides, we identified enrichment in the Wnt signaling pathway, also enriched in differential 5mC analysis, and known to be involved in opioid-related withdrawal symptoms in mice^20^. Another interesting, enriched pathway was the inhibition of voltage-gated Ca^2+^ channels via inhibition of G protein-gated potassium channels. Voltage-gated Ca^2+^ channels are involved in the release of pain neurotransmitters^28^ and are a target of pain-relieving drugs. For co-hydroxymethylation, we identified modules enriched for class B/2 (secretin family receptor), which was also identified in a co-expression analysis of maternal exposure to oxycodone^29^. Further, we identified the IGF1R signaling cascade and insulin receptor signaling cascade. It is known that opioids affect glucose homeostasis^30^, being suggested a role of opioids in the inhibition of insulin receptor during morphine use^31^. We also performed a PPI analysis to identify functional gene networks based on the 5mC and 5hmC modules identified. We found evidence of co-expression in genes involved in neurogenesis pathways, cell differentiation, and neuronal development for both 5mC and 5hmC. Previous studies have shown that opioids decrease neurogenesis by inhibiting cell division, mainly by blocking the S phase^32^. We detected one differential 5hmC annotated gene in two 5hmC modules, *CCT2* gene in navajowhite3 and rosybrown1, while no annotated genes were observed for 5mC. *CCT2* is a chaperonin with a role in hypoxia in colorectal cancer^33^. Our findings revealed epigenetically-dysregulated gene networks in OUD involving the Wnt, immune, and pain signaling pathways as well as neurogenesis.

To evaluate the potential effects of genetic background in the differential 5mC and 5hmC marks and modules, we conducted a GWAS enrichment analysis. We found significant enrichment of OUD-linked 5hmC modules with schizophrenia and bipolar affective disorders, as in the differential 5hmC CpGs, but also with cognitive function domains, depression, and substance use disorders.

To better understand the impact of 5mC and 5hmC alterations on gene expression, we conducted a differential gene expression analysis followed by integration with 5mC and 5hmC data. Differential analysis of RNA sequencing data from bulk tissue identified the Hemoglobin Subunit Beta (*HBB*) gene as associated with OUD. Hemoglobin acts as an oxygen-storage molecule in hypoxia, suggested to be induced by opioids^34, 35^, suggesting an increase in brain hypoxia in the OFC of OUD individuals. Correlation analysis between 5mC/5hmC and gene expression levels detected an overlap in 64 differential CpG sites for 5mC and 257 for 5hmC. In addition, a higher correlation was detected for 5hmC CpGs in the *TMIGD3* (Transmembrane and Immunoglobulin Domain Containing 3) gene. *TMIGD3* can act as a repressor of NF-κB^36^, an essential protein that regulates inflammation and is altered in individuals with OUD^37, 38^. These results suggest a higher impact of 5hmC than 5mC on gene regulation in OUD.

This study has several strengths. It is the first study assessing 5hmC in parallel with 5mC in OUD. Most studies have used bulk tissue, where molecular changes may be obscured, as also evident in our bulk gene expression results. We performed FANS-sorted neuronal nuclei, which allowed us to conduct the 5mC/5hmC mapping of the human OFC in a cell-type-specific manner. Leveraging RRoxBS resulted in three times more CpG coverage than the latest and most commonly used microarray technology (Infinium MethylationEPIC Array). We also conducted RNA sequencing analysis for the same subjects and integrated it with 5mC and 5hmC data to evaluate the effects of these epigenetic modifications on gene transcription. 5mC and 5hmC patterns were assessed using two analytical approaches: CpG site and co-methylation analysis, which allow us not only single CpG loci associations, but also epigenetically-regulated gene networks. We also integrated our epigenetic findings with GWAS data showing an association of OUD-associated 5hmC genes and modules with psychiatric disorders, including substance use disorders.

The study is limited by the small sample size, but it is comparable to similar work in the human postmortem brain. All OUD subjects were comorbid with PTSD. We addressed this issue by adding PTSD as a covariate in the model (PTSD is present in 50% of the non-OUD group) and replicated previous work assessing 5mC in the OFC of heroin abusers^11^. RNA sequencing data was conducted in bulk tissue, which may impede our ability to identify OUD DEGs and directly evaluate the impact of differential 5mC/5hmC on gene expression in a cell-type-specific manner. Another limitation is the sole inclusion of males, mostly of European ancestry. Future work will expand to female subjects and other ancestries to identify potential sex- and population-specific effects. Further, additional comorbidities (e.g., MDD and other SUDs) should be also examined. This study evaluated 5mC and 5hmC at CpG sites; future studies should evaluate the role of 5mC and 5hmC in non-CpG sites in the context of OUD, which have been suggested to play a crucial role in the development of neuropsychiatric diseases^39^. Given the enrichment of differential 5mC and 5hmC genes and modules in GWAS studies, future work should evaluate potential methylation/hydroxymethylation quantitative trait loci to assess whether differential 5mC/5hmC is influenced by genetic background. Though the enrichment of GWAS signals may be indicative of potential causal effects of the OUD-associated 5mC/5hmC marks, research in model organisms (i.e., animal models, human induced pluripotent stem cells) could help confirm whether the identified marks are the cause or consequence of OUD.

In summary, our findings identified 5mC and 5hmC dysregulation in OFC neurons from individuals with OUD. The results suggest that 5hmC plays an important role in OUD, as shown by the magnitude and functionality of differential marks and networks, as well as its potential impact on gene expression patterns. We have identified clinically relevant pathways, such as NOTCH signaling processes, Wnt signaling, and G protein signaling pathways, as well as enrichment for psychiatric domains, including substance use disorders. Lastly, multiple gene-drug interactions between differential 5mC/5hmC and opioids were identified, revealing targets of well-known and potentially novel OUD treatments. Our study supports the important role of 5hmC on OUD and demonstrates a multi-omic dysregulation of OUD in the human OFC, identifying well-known and novel targets that may inform prognostic and treatment efforts in the future.

## Methods

### Sample description

Postmortem human brain specimens were obtained from the National Post-Traumatic Stress Disorder (PTSD) Brain Bank^40^ (NPBB), a brain tissue repository at the U.S. Department of Veterans Affairs (VA). In addition, brain tissue samples were collected from the orbitofrontal cortex (OFC; Brodmann Area 11). **Table 1** shows the demographics and clinical characteristics of the study cohort. Diagnoses were conducted using two approaches, antemortem assessment protocol for antemortem donors and postmortem diagnostic assessment for postmortem donors as described by Friedman and colleagues^40^.

### Fluorescence-Activated Nuclei Sorting

For each specimen, OFC tissue (100-200 mg) was lysed in homogenization buffer (0.1% Triton, 0.32 M sucrose, 5 mM CaCl2, 3 mM MgCl2, 10 mM Tris-HCl) on ice in a glass Dounce homogenizer. Homogenized samples were filtered through a 40 µM cell strainer, loaded onto a 1.8 M sucrose cushion, and ultracentrifuged (SW-41 rotor, Beckman Coulter, Brea, California, USA) at 24,000 rpm for 1 hour at 4°C. Nuclei were resuspended in 0.5% bovine serum albumin and stained for 30 min at 4°C with Anti-NeuN-PE (Millipore-Sigma, FCMAB317PE). Before sorting, 4′,6-diamidino-2-phenylindole (DAPI) was added as a nuclei label, and samples were again filtered through a 40 µm strainer. FANS procedures were carried out at the Icahn School of Medicine Flow Cytometry CoRE on a BD 5-laser cell sorting system.

### DNA extraction

Between 0.5-1 M NeuN+ nuclei were collected via FANS and processed for DNA extraction. First, sorted nuclei were pelleted by centrifugation at 1500 × g for 15 min at 4°C. Next, the supernatant was aspirated down to 500 µL and 50 µL proteinase K (Cat. #69504, Qiagen, Valencia, CA) and 20 mg/mL RNAse A (Cat. #12091021; Thermo-Fischer, Waltham, MA). Next, samples were processed according to the DNeasy Blood and Tissue Kit (Cat. #69504, Qiagen) manufacturer’s protocol. Finally, eluted samples were further concentrated to 20 µL final volume with the Zymo Genomic DNA Clean and Concentrator-10 kit (Cat. #D4010, Zymo Inc., Irving CA) and stored at -80°C.

### Reduced Representation Oxidative Bisulfite Sequencing (RRoxBS)

RRoxBS was carried out at the Weill Cornell Epigenomics Core (New York, NY). Briefly, two libraries were prepared from 400 ng of DNA to examine methylation and hydroxymethylation at CpG dinucleotides in each subject using the NuGEN Ovation RRoxBS Methyl-Seq library preparation kit. One library underwent bisulfite treatment to convert unmethylated cytosines to uracils, while the second library underwent oxidation prior to bisulfite sequencing (oxBS) to convert both unmethylated and hydroxymethylated cytosines to uracils. Prepared libraries were multiplexed and pooled for single-end 1 × 50 bp sequencing to a mean depth of 42.7 +/- 1.5 (µ +/- SEM) million reads per library on an S4 flow cell using the Illumina NovaSeq6000 system.

### Bioinformatic analysis

The Bismark bisulfite read mapper^41^ was used to map sequencing reads and detect bisulfite treatment of converted and unconverted cytosines. Beta values (% unconverted cytosines) were calculated for 5mC and 5hmC. For all CpGs, 5hmC values were calculated as the difference in methylation at each CpG between oxBS and BS libraries (e.g., if β=100% in the BS library and β=60% in the oxBS library, then 5hmC=40%). Analyzed CpGs were filtered for a minimum of 10x coverage in all subjects and were normalized for coverage variability.

Annotation of CpGs overlapping with neuronal active enhancer regions (H3K27ac) was performed using previously published ChIP-seq datasets^42^ generated from glutamatergic and GABAergic cell types of the human orbitofrontal cortex^42^. In addition, non-neuronal oligodendrocyte enhancers were taken from Nott et al.^43^. Cell type-specific gene markers utilized for enrichment analysis were derived from McKenzie et al.^44^. Regions with total differential methylation (tmC=5mC+5hmC) between NeuN+ and NeuN-populations (>50% difference in tmC) of the human prefrontal cortex were utilized from Rizzardi et al.^17^ to compare with our NeuN+ dataset.

### Differential analysis of methylation and hydroxymethylation

Differential analysis of 5mC and 5hmC was performed at single-CpG resolution with the methylkit R package^45^, using logistic regression with correction for overdispersion and chi-squared significance testing and benchmarked for the best balance of sensitivity and specificity^46^. In addition, we evaluated these epigenetic marks summed over neuronal H3K27ac regions with ≥3 CpGs at ≥10X coverage. Age, ancestry, cigarette smoking, and PTSD were included as *a priori* covariates. 5mC and 5hmC were analyzed separately. Significance was determined as false discovery rate-adjusted p-value<0.05. To investigate functional gene sets enriched for differential CpGs, gene ontology analysis was carried out using the methylGSA Bioconductor package^47^ with gene lists analyzed from the NCATS BioPlanet, Gene Ontology Consortium, and Kyoto Encyclopedia of Genes and Genomes (KEGG) databases. Furthermore, enrichment of differentially methylated markers for genomic regions and features were evaluated using one-sided Fisher’s exact tests.

### Co-methylation analysis

Co-methylation analysis was performed for 5mC and 5hmC, separately, using the WGCNA R package^48^. QC was performed to remove sites with missing values. 1,835,925 and 1,638,378 sites were kept in the co-methylation analysis of 5mC and 5hmC, respectively. Age, ancestry, PTSD, and smoking status were included as covariates using the empiricalBayesLM function from the WGCNA package. The blockwiseModules function was used to conduct the module detection. Soft-threshold power was set as 5 and TOMType as signed for both analyses. The eigengene calculated for each module was used for the correlation analysis with OUD. Correlation greater than |±0.4| and p-value<0.05 was considered significant. AmiGO^49^ was used for enrichment analysis, considering Gene Ontology (GO) for biological processes and Reactome pathways.

The nearest gene IDs were submitted to protein-protein interaction (PPI) using STRING^50^. We selected genes from the top 20 biological processes GO terms after PPI analysis to reduce the number of nodes. For the thistle1 module, we used genes from the top 10 biological processes GO terms. The parameters were set to show evidence based on co-expression data, applying 0.9 of the minimum required interaction score. We used two approaches to assess the enrichment of differential 5mC and 5hmC genes. First, we calculated a rate representing [Number of observed differential sites in the module]/[Number of expected differential sites]^19^. Then we calculated the enrichment p-value using the fisher.test function.

### RNA sequencing

RNA from bulk-tissue medial OFC (BA 11) was extracted for all samples as previously described^51^. Briefly, 20mg of tissue was isolated using the RNeasy Mini Kit (Qiagen) with genomic DNA elimination. RNA integrity and concentration were measured using a Bioanalyzer (Agilent) and rRNA was depleted using Ribo-Zero Gold Kit (Illumina). Libraries were constructed using the SMARTer® Stranded RNA-seq Kit (Takara Bio) and sequenced at 75 bp paired-end on an Illumina HiSeq4000.

### Differential gene expression

FASTQ files were mapped and annotated as previously described^51^. Differentially expressed genes (DEGs) were calculated using the DESeq2 package in R^52^. That statistical framework enabled the calculation of log2 fold-change values (log2FC) for each gene in the PTSD+OUD and PTSD+CON raw count data. This model considered the following covariates to measure the association with opioid use: age, RNA integrity number (RIN), PTSD diagnosis, and smoking status. Genes with zero expression in all samples of a group were dropped. Differentially expressed genes were defined by an FDR-adjusted p-value<0.05 (Benjamini-Hochberg).

### Correlation analysis between gene expression and 5mC/5hmC

Pearson correlation between mRNA analysis and 5mC/5hmC data was calculated using the MatrixeQTL package^53^. Correlation analysis was performed in *cis* and *trans*. The raw count data from the RNAseq analysis was normalized by variance stabilization transformation, and the matrix was used for correlation. We adjusted the correlations for age, RIN, PTSD, and smoking status. Significance was defined by an FDR-adjusted p-value<0.05.

### GWAS enrichment analysis

Genome-wide association enrichment signals analysis was performed using the online website FUMA^54^. First, the Ensembl IDs were transformed to Entrez IDs using David^55^ and submitted to FUMA’s GENE2Function web tool. Once the enrichment analysis was generated, the results were merged with the domains and traits reported by the GWAS Atlas using in-house scripts.

### Drug interaction analysis

Drug interaction analysis was conducted for the annotated genes of the differential 5mC and 5hmC CpGs using the Drug Gene Interaction Database (DGIdb; https://www.dgidb.org)^56^. From the gene-drug interactions identified, we further evaluated those in 15 opiates: apomorphine, codeine, diacetylmorphine, hydrocodone, methadone, morphine, oxycodone, oxymorphone, tramadol, methadone, propoxyphene, fentanyl, hydromorphone, heroin, and levorphanol. In addition to opioids, we also evaluated other drugs that showed interactions with the same annotated genes.

## Data Availability

Summary-level data produced in the present work are included in the Supplementary materials. All data produced are available upon reasonable request.

## Data availability

Summary statistics are available in Supplementary Tables.

## Acknowledgments

This work is supported by the U.S. Department of Veterans Affairs via 1IK2CX002095-01A1 (J.L.M.O.) and NIDA R21DA050160 (J.L.M.O.). We thank Dr. Sarah Beck for the editing of the manuscript.

## Members of the Traumatic Stress Brain Research Group

Victor E. Alvarez, MD; David Benedek, MD; Alicia Che, PhD; Dianne A. Cruz, MS; David A. Davis, PhD Matthew J. Girgenti; PhD, Ellen Hoffman, MD, PhD; Paul E. Holtzheimer, MD; Bertrand R. Huber, MD, PhD; Alfred Kaye, MD, PhD; John H. Krystal, MD; Adam T. Labadorf, PhD; Terence M. Keane, PhD; Mark W. Logue, PhD; Ann McKee, MD; Brian Marx, PhD; Deborah Mash, MD; Mark W. Miller, PhD; Crystal Noller, PhD; Janitza Montalvo-Ortiz, PhD; William K. Scott, PhD; Paula Schnurr, PhD; Thor Stein, MD, PhD; Robert Ursano, MD; Douglas E. Williamson, PhD; Erika J. Wolf, PhD, Keith A. Young, PhD

## Disclosures

The following competing interests for JHK: 1) Consultant: Note: –The Individual Consultant Agreements listed below are less than $10,000 per year: AstraZeneca Pharmaceuticals; Biogen, Idec, MA; Biomedisyn Corporation; Bionomics, Limited (Australia); Concert Pharmaceuticals, Inc.; Heptares Therapeutics, Limited (UK); Janssen Research & Development; L.E.K. Consulting; Otsuka America Pharmaceutical, Inc.; Spring Care, Inc.; Sunovion Pharmaceuticals, Inc.; Takeda Industries; Taisho Pharmaceutical Co., Ltd; Scientific Advisory Board; Bioasis Technologies, Inc.; Biohaven Pharmaceuticals; Blackthorn Therapeutics, Inc.; Broad Institute of MIT and Harvard; Cadent Therapeutics; Lohocla Research Corporation; Pfizer Pharmaceuticals ; Stanley Center for Psychiatric Research at the Broad Institute; 2) Stock: ArRETT Neuroscience, Inc.; Blackthorn Therapeutics, Inc.; Biohaven Pharmaceuticals Medical Sciences; Spring Care, Inc. Stock Options: Biohaven PharmaceuticalsMedical Sciences; 3) Income Greater than $10,000: Editorial BoardEditor –Biological Psychiatry; Patents and Inventions: Seibyl JP, Krystal JH, Charney DS. Dopamine and noradrenergic reuptake inhibitors in treatment of schizophrenia. US Patent #:5,447,948. September 5, 1995; Vladimir, Coric, Krystal, John H, Sanacora, Gerard – Glutamate Modulating Agents in the Treatment of Mental Disorders US Patent No. 8,778,979 B2 Patent Issue Date: July 15, 2014. US Patent Application No. 15/695,164: Filing Date: 09/05/2017; Charney D, Krystal JH, Manji H, Matthew S, Zarate C., -Intranasal Administration of Ketamine to Treat Depression United States Application No. 14/197,767 filed on March 5, 2014; United States application or Patent Cooperation Treaty (PCT) International application No. 14/306,382 filed on June 17, 2014; Zarate, C, Charney, DS, Manji, HK, Mathew, Sanjay J, Krystal, JH, Department of Veterans Affairs “Methods for Treating Suicidal Ideation”, Patent Application No. 14/197.767 filed on March 5, 2014 by Yale University Office of Cooperative Research; Arias A, Petrakis I, Krystal JH. –Composition and methods to treat addiction; Provisional Use Patent Application no.61/973/961. April 2, 2014. Filed by Yale University Office of Cooperative Research; Chekroud, A., Gueorguieva, R., & Krystal, JH. “Treatment Selection for Major Depressive Disorder” [filing date 3^rd^ June 2016, USPTO docket number Y0087.70116US00]. Provisional patent submission by Yale University; Yoon G, Petrakis I, Krystal JH. – Compounds, Compositions and Methods for Treating or Preventing Depression and Other Diseases. U. S. Provisional Patent Application No. 62/444,552, filed on January10, 2017 by Yale University Office of Cooperative Research OCR 7088 US01; Abdallah, C, Krystal, JH, Duman, R, Sanacora, G. Combination Therapy for Treating or Preventing Depression or Other Mood Diseases. U.S. Provisional Patent Application No. 047162-7177P1 (00754) filed on August 20, 2018 by Yale University Office of Cooperative Research OCR 7451 US01. Dr. Gelernter is named as an inventor on PCT patent application #15/878,640 entitled: “Genotype-guided dosing of opioid agonists,” filed January 24, 2018 and issued on January 26, 2021 as U.S. Patent No. 10,900,082. JG is paid for editorial work for the journal “Complex Psychiatry.” Other co-authors declare no competing interests.

## Author contribution

G.R. and S.T.N conducted the analyses and drafted the manuscript, Y.H. conceived and designed the study and revised the manuscript, J.J.M.M. performed GWAS enrichment and correlation analyses between gene expression and 5mC/5hmC data and assisted in manuscript writing, Methods section, M.J.G. and J.W. performed the RNA-seq analysis and assisted in manuscript writing, Methods section, J.H.K. and J.G. provided feedback and revised the manuscript. J.L.M.O. conceived, designed, and coordinated the study and supervised the manuscript preparation and revision. All authors contributed to and approved the final manuscript.

## Supplementary Tables

1 Differential CpG Methylation (FDR-adjusted q<0.05)

2 Differential CpG Hydroxymethylation (FDR-adjusted q<0.05)

3 Genomic Region and Feature Enrichment Analysis

4 Gene Ontology for Differentially Methylated Genes

5 Gene Ontology for Differentially Hydroxymethylated Genes

6 Overlapping Differentially Methylated Genes between OUD and Heroin Overdose

7 Gene Ontology for Differentially Methylated Genes Shared Between OUD and Heroin Overdose

8 Gene Ontology for 5mC Co-Methylation

9 Gene Ontology for 5hmC Co-Methylation

10 Reactome Pathways of 5mC and 5hmC Modules

11 Differential Expression Genes

12 Methylated sites correlated with gene expression

13 Hydroxymethylated sites correlated with gene expression

14 GWAS for 5hmC

15 GWAS for Co-Methylated Modules

## Supplementary Figures

**Supplementary Figure 1. Representative FANS gating**. FANS of single nuclei. Nuclei stained with NeuN-PE conjugated antibody were filtered through a 40-μm cell strainer and loaded onto a custom FACS ARIA II flow sorter (Becton Dickinson) equipped with a forward scatter photomultiplier tube. (A) Particles smaller than nuclei were first eliminated with an area plot of forward scatter (FSC-A) vs. side scatter (SSC-A). (B-C) Plots of area vs. width in the forward and side scatter channels, respectively, are used for doublet discrimination with gating to exclude aggregates of 2 or more nuclei. (D) Remaining nuclei were then gated in the violet wavelength while excluding the remaining doublet and triplet signals. (E) The final gating panel captured NeuN+ neuronal nuclei using the yellow emission spectra. NeuN+ nuclei represented ~50-60% of DAPI+ nuclei.

**Supplementary Figure 2**. A) Principal component analysis to identify outliers. B) Distribution of 5mC CpGs in the gene region. C) Distribution of 5hmC CpGs in the gene region.

**Supplementary Figure 3. Exploring 5mC and 5hmC co-methylation results**. A) Module membership vs. gene significance in 5mC co-methylation analysis. B) Module membership vs. gene significance in 5hmC co-methylation analysis. C) Number of observed CpGs for modules with rate Observed/Expected>=2 for 5mC (left) and 5hmC (right).

**Supplementary Figure 4. Genome-wide association enrichment signals analysis**. A) GWAS enrichment analysis of the annotated genes for 5hmC differential CpGs. B) GWAS enrichment analysis for co-methylated and co-hydroxymethylated modules.

## Notes

### Author Declarations

VA Institutional Review Board.

### Summary of Updates

Update the figures

## References

1. French R, Aronowitz SV, Brooks Carthon JM, Schmidt HD, Compton P. Interventions for hospitalized medical and surgical patients with opioid use disorder: A systematic review. Subst Abus 43, 495–507 (2022).

2. Barton SJ, et al. In Epigenomic Studies, Including Cell-Type Adjustments in Regression Models Can Introduce Multicollinearity, Resulting in Apparent Reversal of Direction of Association. Front Genet 10, 816 (2019).

3. A time of crisis for the opioid epidemic in the USA. Lancet 398, 277–277 (2021).

4. Nestler EJ. Historical review: Molecular and cellular mechanisms of opiate and cocaine addiction. Trends Pharmacol Sci 25, 210–218 (2004).

5. Redmond DE, Jr., Krystal JH. Multiple mechanisms of withdrawal from opioid drugs. Annu Rev Neurosci 7, 443–478 (1984).

6. Olausson P, Jentsch JD, Krueger DD, Tronson NC, Nairn AC, Taylor JR. Orbitofrontal cortex and cognitive-motivational impairments in psychostimulant addiction: evidence from experiments in the non-human primate. Ann N Y Acad Sci 1121, 610–638 (2007).

7. Lasseter HC, Ramirez DR, Xie X, Fuchs RA. Involvement of the lateral orbitofrontal cortex in drug context-induced reinstatement of cocaine-seeking behavior in rats. Eur J Neurosci 30, 1370–1381 (2009).

8. Schoenbaum G, Shaham Y. The role of orbitofrontal cortex in drug addiction: a review of preclinical studies. Biol Psychiatry 63, 256–262 (2008).

9. Fuchs RA, Evans KA, Parker MP, See RE. Differential involvement of orbitofrontal cortex subregions in conditioned cue-induced and cocaine-primed reinstatement of cocaine seeking in rats. J Neurosci 24, 6600–6610 (2004).

10. Browne CJ, Godino A, Salery M, Nestler EJ. Epigenetic Mechanisms of Opioid Addiction. Biol Psychiatry 87, 22–33 (2020).

11. Kozlenkov A, et al. DNA Methylation Profiling of Human Prefrontal Cortex Neurons in Heroin Users Shows Significant Difference between Genomic Contexts of Hyper- and Hypomethylation and a Younger Epigenetic Age. Genes (Basel) 8, (2017).

12. Wang T, et al. Genome-wide DNA hydroxymethylation changes are associated with neurodevelopmental genes in the developing human cerebellum. Hum Mol Genet 21, 5500–5510 (2012).

13. Cui XL, et al. A human tissue map of 5-hydroxymethylcytosines exhibits tissue specificity through gene and enhancer modulation. Nat Commun 11, 6161 (2020).

14. Gross JA, et al. Gene-body 5-hydroxymethylation is associated with gene expression changes in the prefrontal cortex of depressed individuals. Transl Psychiatry 7, e1119 (2017).

15. Clark SL, et al. Dual methylation and hydroxymethylation study of alcohol use disorder. Addict Biol 27, e13114 (2022).

16. Barrow TM, et al. The effect of morphine upon DNA methylation in ten regions of the rat brain. Epigenetics 12, 1038–1047 (2017).

17. Rizzardi LF, et al. Neuronal brain-region-specific DNA methylation and chromatin accessibility are associated with neuropsychiatric trait heritability. Nat Neurosci 22, 307–316 (2019).

18. Creyghton MP, et al. Histone H3K27ac separates active from poised enhancers and predicts developmental state. Proc Natl Acad Sci U S A 107, 21931–21936 (2010).

19. Wang F, Xu H, Zhao H, Gelernter J, Zhang H. DNA co-methylation modules in postmortem prefrontal cortex tissues of European Australians with alcohol use disorders. Sci Rep 6, 19430 (2016).

20. Wu M, et al. Wnt signaling contributes to withdrawal symptoms from opioid receptor activation induced by morphine exposure or chronic inflammation. Pain 161, 532–544 (2020).

21. Williams JT, et al. Regulation of mu-opioid receptors: desensitization, phosphorylation, internalization, and tolerance. Pharmacol Rev 65, 223–254 (2013).

22. Li D, et al. Association of the calcyon neuron-specific vesicular protein gene (CALY) with adolescent smoking initiation in China and California. Am J Epidemiol 173, 1039–1048 (2011).

23. Ge X, Qiu Y, Loh HH, Law PY. GRIN1 regulates micro-opioid receptor activities by tethering the receptor and G protein in the lipid raft. J Biol Chem 284, 36521–36534 (2009).

24. Boroumand F, Mahmoudinasab H, Saadat M. Association of the SOD2 (rs2758339 and rs5746136) polymorphisms with the risk of heroin dependency and the SOD2 expression levels. Gene 649, 27–31 (2018).

25. Munoz B, Fritz BM, Yin F, Atwood BK. HCN1 channels mediate mu opioid receptor long-term depression at insular cortex inputs to the dorsal striatum. bioRxiv preprint, (2022).

26. Zhang J, et al. Hypermethylation in the promoter region of the ADRA1A gene is associated with opioid use disorder in Han Chinese. Brain Res 1793, 148050 (2022).

27. Sanna MD, Borgonetti V, Galeotti N. mu Opioid Receptor-Triggered Notch-1 Activation Contributes to Morphine Tolerance: Role of Neuron-Glia Communication. Mol Neurobiol 57, 331–345 (2020).

28. Lee S. Pharmacological Inhibition of Voltage-gated Ca(2+) Channels for Chronic Pain Relief. Curr Neuropharmacol 11, 606–620 (2013).

29. Green MT, et al. Maternal oxycodone treatment causes pathophysiological changes in the mouse placenta. Placenta 100, 96–110 (2020).

30. Toorie AM, Vassoler FM, Qu F, Slonim D, Schonhoff CM, Byrnes EM. Intergenerational effects of preconception opioids on glucose homeostasis and hepatic transcription in adult male rats. Sci Rep 12, 1599 (2022).

31. Li Y, et al. Morphine induces desensitization of insulin receptor signaling. Mol Cell Biol 23, 6255–6266 (2003).

32. Salarinasab S, et al. Interaction of opioid with insulin/IGFs signaling in Alzheimer’s disease. J Mol Neurosci 70, 819–834 (2020).

33. Park SH, et al. Activating CCT2 triggers Gli-1 activation during hypoxic condition in colorectal cancer. Oncogene 39, 136–150 (2020).

34. Kiyatkin EA. Respiratory depression and brain hypoxia induced by opioid drugs: Morphine, oxycodone, heroin, and fentanyl. Neuropharmacology 151, 219–226 (2019).

35. Hayen A, et al. Opioid suppression of conditioned anticipatory brain responses to breathlessness. Neuroimage 150, 383–394 (2017).

36. Iyer SV, et al. Genome-wide RNAi screening identifies TMIGD3 isoform1 as a suppressor of NF-kappaB and osteosarcoma progression. Nat Commun 7, 13561 (2016).

37. Seney ML, et al. Transcriptional Alterations in Dorsolateral Prefrontal Cortex and Nucleus Accumbens Implicate Neuroinflammation and Synaptic Remodeling in Opioid Use Disorder. Biol Psychiatry 90, 550–562 (2021).

38. Bryant BM, Eaton E, Li L. A Systematic Review of Opioid Use Disorder and Related Biomarkers. Front Psychiatry 12, 708283 (2021).

39. Jang HS, Shin WJ, Lee JE, Do JT. CpG and Non-CpG Methylation in Epigenetic Gene Regulation and Brain Function. Genes (Basel) 8, (2017).

40. Friedman MJ, et al. VA’s National PTSD Brain Bank: a National Resource for Research. Curr Psychiatry Rep 19, 73 (2017).

41. Krueger F, Andrews SR. Bismark: a flexible aligner and methylation caller for Bisulfite-Seq applications. Bioinformatics 27, 1571–1572 (2011).

42. Kozlenkov A, et al. A unique role for DNA (hydroxy)methylation in epigenetic regulation of human inhibitory neurons. Sci Adv 4, eaau6190 (2018).

43. Nott A, et al. Brain cell type-specific enhancer-promoter interactome maps and disease-risk association. Science 366, 1134–1139 (2019).

44. McKenzie AT, et al. Brain Cell Type Specific Gene Expression and Co-expression Network Architectures. Sci Rep 8, 8868 (2018).

45. Akalin A, et al. methylKit: a comprehensive R package for the analysis of genome-wide DNA methylation profiles. Genome Biol 13, R87 (2012).

46. Wreczycka K, Gosdschan A, Yusuf D, Gruning B, Assenov Y, Akalin A. Strategies for analyzing bisulfite sequencing data. J Biotechnol 261, 105–115 (2017).

47. Ren X, Kuan PF. methylGSA: a Bioconductor package and Shiny app for DNA methylation data length bias adjustment in gene set testing. Bioinformatics 35, 1958–1959 (2019).

48. Langfelder P, Horvath S. WGCNA: an R package for weighted correlation network analysis. BMC Bioinformatics 9, 559 (2008).

49. Carbon S, et al. AmiGO: online access to ontology and annotation data. Bioinformatics 25, 288–289 (2009).

50. Szklarczyk D, et al. STRING v11: protein-protein association networks with increased coverage, supporting functional discovery in genome-wide experimental datasets. Nucleic Acids Res 47, D607–D613 (2019).

51. Girgenti MJ, et al. Transcriptomic organization of the human brain in post-traumatic stress disorder. Nat Neurosci 24, 24–33 (2021).

52. Love MI, Huber W, Anders S. Moderated estimation of fold change and dispersion for RNA-seq data with DESeq2. Genome Biol 15, 550 (2014).

53. Shabalin AA. Matrix eQTL: ultra fast eQTL analysis via large matrix operations. Bioinformatics 28, 1353–1358 (2012).

54. Watanabe K, Taskesen E, van Bochoven A, Posthuma D. Functional mapping and annotation of genetic associations with FUMA. Nat Commun 8, 1826 (2017).

55. Huang DW, et al. The DAVID Gene Functional Classification Tool: a novel biological module-centric algorithm to functionally analyze large gene lists. Genome Biol 8, R183 (2007).

56. Freshour SL, et al. Integration of the Drug-Gene Interaction Database (DGIdb 4.0) with open crowdsource efforts. Nucleic Acids Res 49, D1144–D1151 (2021).

