## Supplementary Figure for "Profiling Neuronal Methylome and Hydroxymethylome of Opioid Use Disorder in the Human Orbitofrontal Cortex"

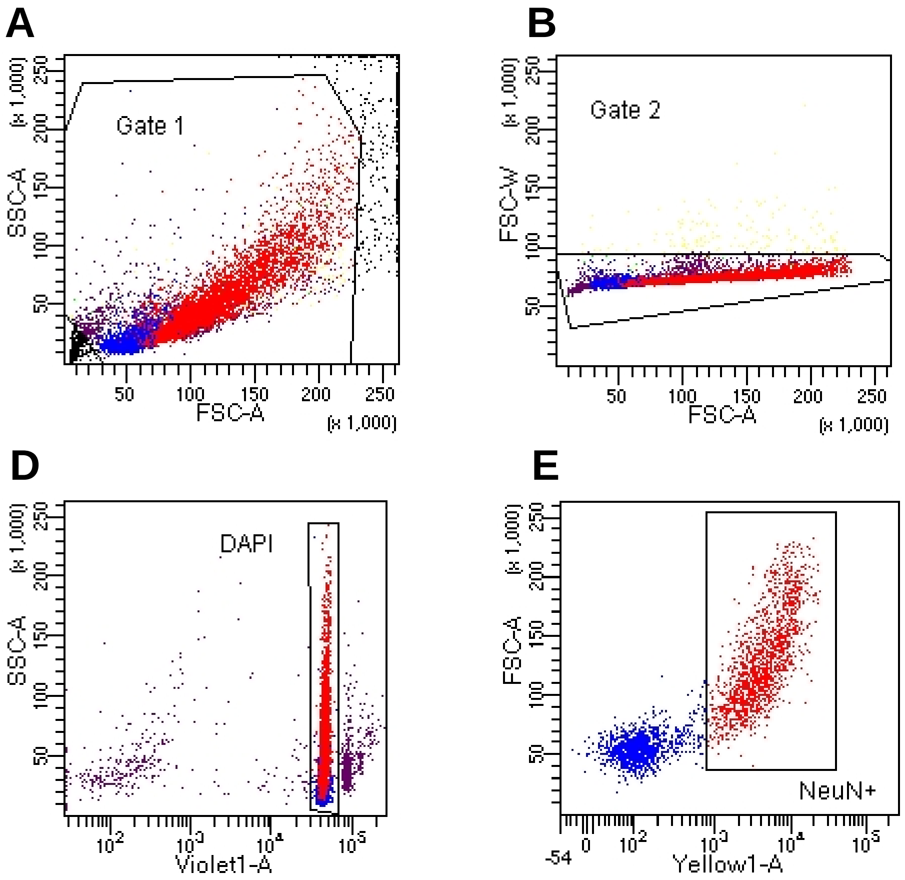


F

Tube: Representative sample

| Population | #Events | %Parent | %Total |
|---|---|---|---|
| All Events | 10,000 | #### | 100.0 |
| Gate 1 | 9,035 | 90.4 | 90.4 |
| Gate 2 | 8,886 | 98.4 | 88.9 |
| Gate 3 | 8,871 | 99.8 | 88.7 |
| DAPI | 8,038 | 90.6 | 80.4 |
| NeuN+ | 4,382 | 54.5 | 43.8 |

**Supplementary Figure 1. Representative FANS gating.** FANS of single nuclei. Nuclei stained with NeuN-PE conjugated antibody were filtered through a 40-µm cell strainer and loaded onto a custom FACS ARIA II flow sorter (Becton Dickinson) equipped with a forward scatter photomultiplier tube. (A) Particles smaller than nuclei were first eliminated with an area plot of forward scatter (FSC-A) vs. side scatter (SSC-A). (B-C) Plots of area vs. width in the forward and side scatter channels, respectively, are used for doublet discrimination with gating to exclude aggregates of 2 or more nuclei. (D) Remaining nuclei were then gated in the violet wavelength while excluding the remaining doublet and triplet signals. (E) The final gating panel captured NeuN+ neuronal nuclei using the yellow emission spectra. NeuN+ nuclei represented ~50-60% of DAPI+ nuclei.

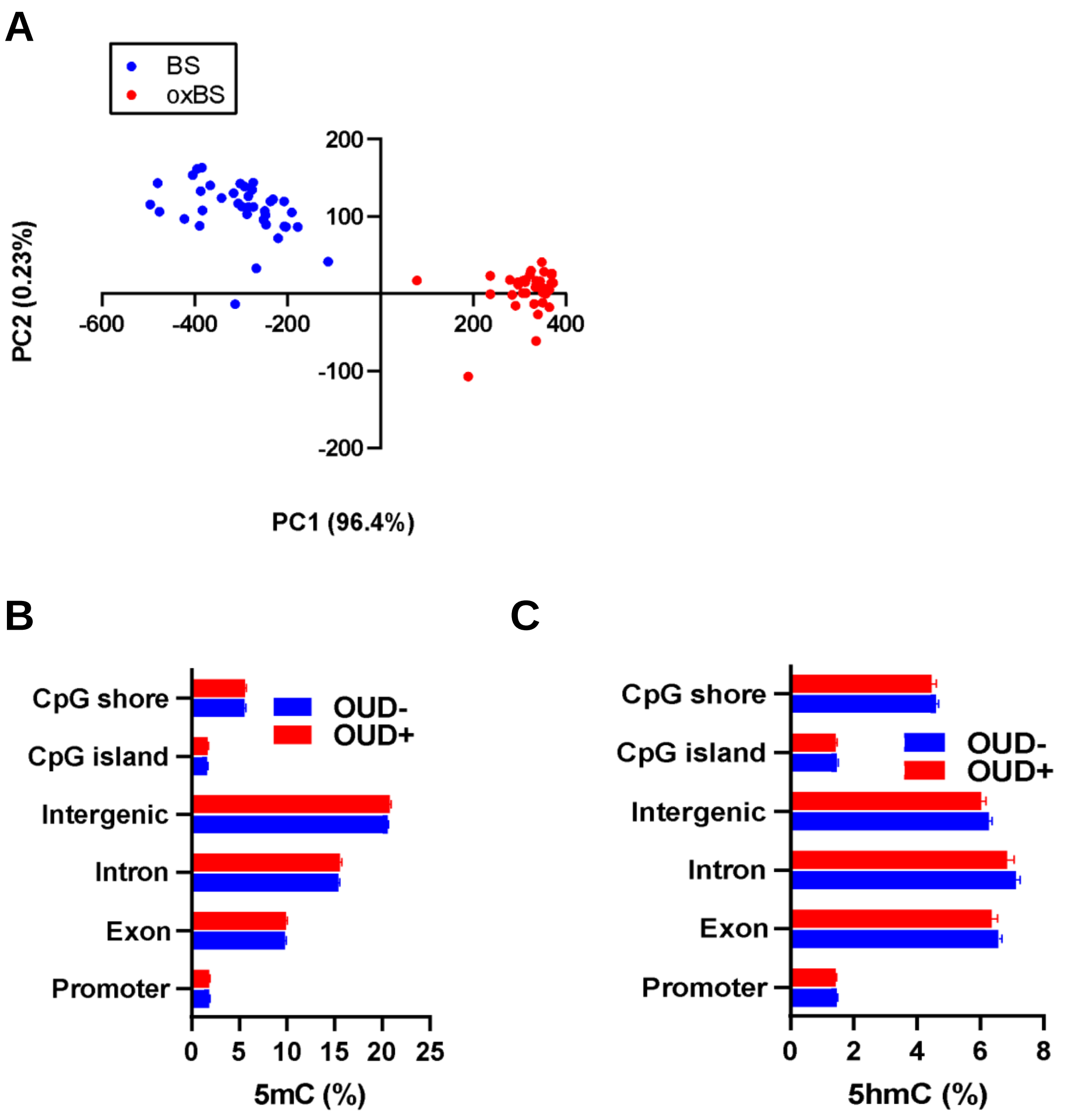


**Supplementary Figure 2.** A) Principal component analysis to identify outliers. B) Distribution of 5mC CpGs in the gene region. C) Distribution of 5hmC CpGs in the gene region.

**A** **Module Membership vs. gene significance 5mC**

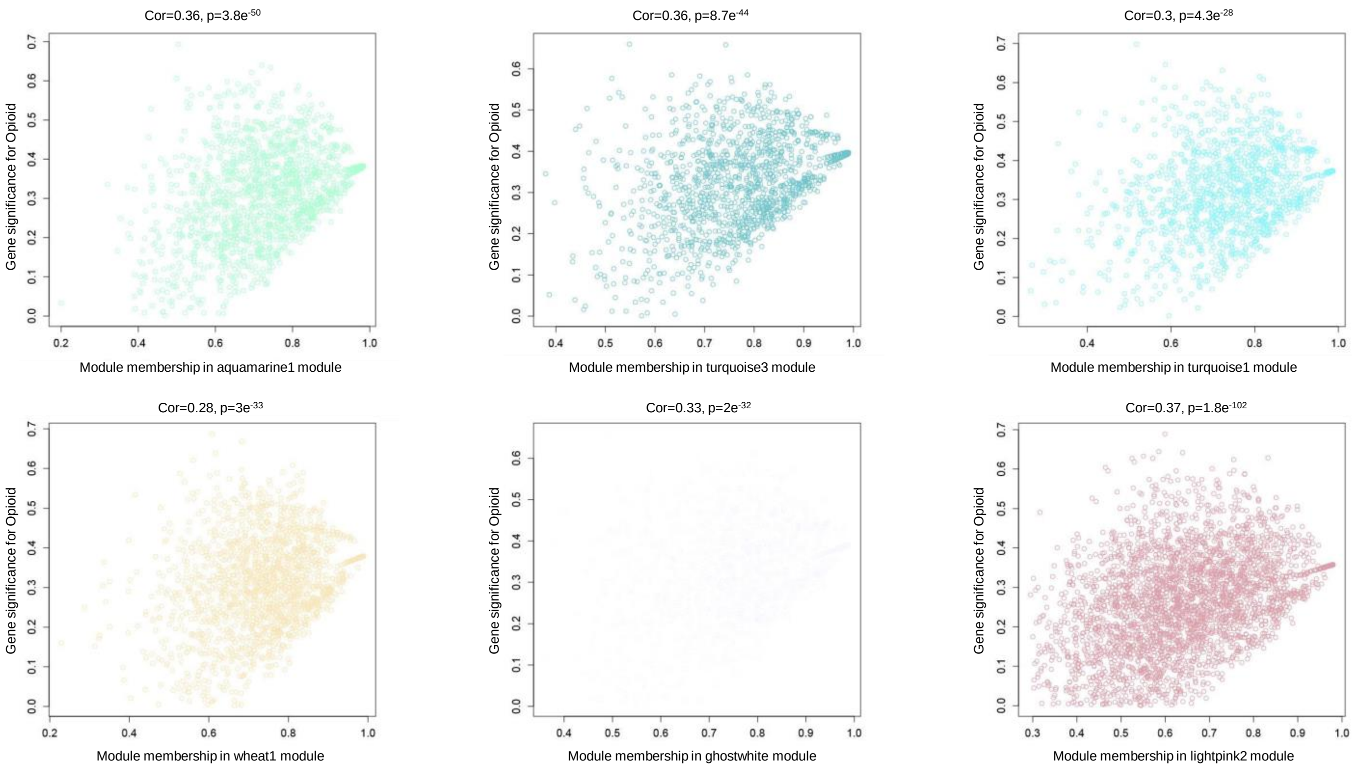


**B** **Module Membership vs. gene significance 5hmC**

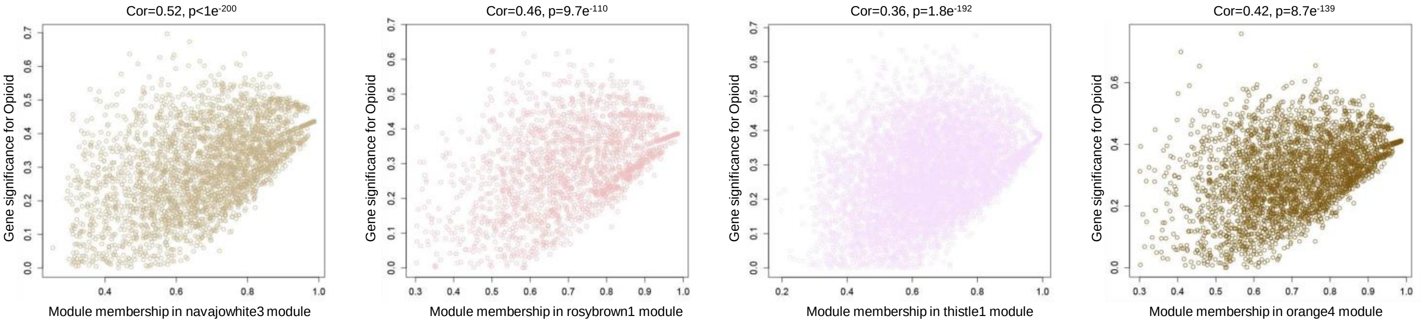


**C** **Distribution of differential sites into modules**

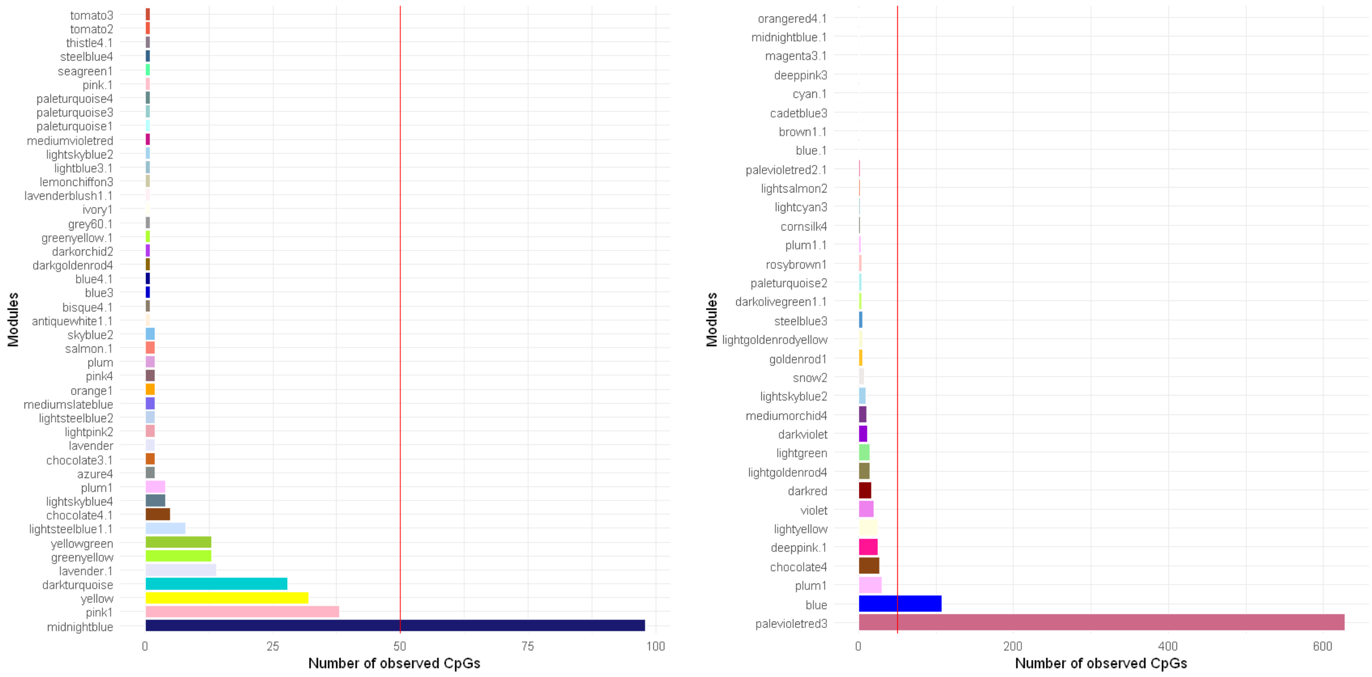


**Supplementary Figure 3.** Exploring 5mC and 5hmC co-methylation results. A) Module membership vs. gene significance in 5mC co-methylation analysis. B) Module membership vs. gene significance in 5hmC co-methylation analysis. C) Number of observed CpGs for modules with rate Observed/Expected>=2 for 5mC (left) and 5hmC (right).

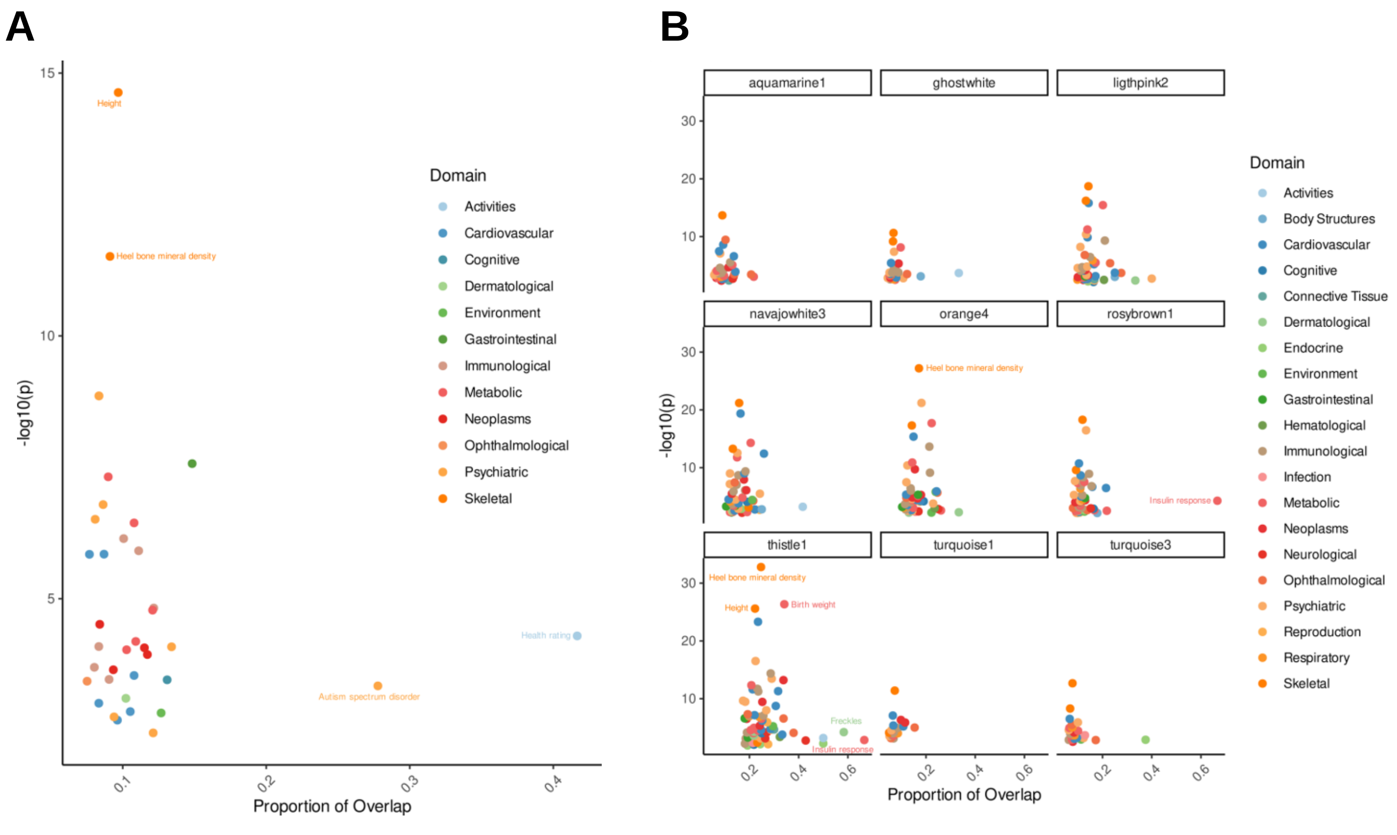


**Supplementary Figure 4.** Genome-wide association enrichment signals analysis. A) GWAS enrichment analysis of the annotated genes for 5hmC differential CpGs. B) GWAS enrichment analysis for co-methylated and co-hydroxymethylated modules.
